# Impact of SARS-CoV-2 on Seasonal Respiratory Viruses: A Tale of Two Large Metropolitan Centers in the United States

**DOI:** 10.1101/2020.10.15.20213371

**Authors:** Amy C. Sherman, Ahmed Babiker, Andrew J. Sieben, Alexander Pyden, James Steinberg, Colleen S. Kraft, Katia Koelle, Sanjat Kanjilal

## Abstract

To assess the impact of the SARS-CoV-2 pandemic on seasonal respiratory viruses, absolute case counts and viral reproductive rates from 2019-2020 were compared against previous seasons. Our findings suggest that the public health measures implemented to reduce SARS-CoV-2 transmission significantly reduced the transmission of other respiratory viruses.

---

The severe acute respiratory syndrome coronavirus-2 (SARS-CoV-2) pandemic has resulted in over 7.4 million cases of coronavirus disease 2019 (COVID-19) and over 210,000 deaths in the United States alone, as of October 7, 2020 [1]. The introduction of SARS-CoV-2 into the northern hemisphere in January 2020 overlapped with the peak circulation of seasonal respiratory viruses that include influenza A and B and respiratory syncytial virus (RSV). Like SARS-CoV-2, these viruses are also spread primarily through droplet and contact transmission routes, with concern that co-circulating respiratory pandemics could further stress our healthcare systems and economy [2, 3].

The danger represented by the unchecked spread of SARS-CoV-2 prompted a massive rollout of population-level non-pharmaceutical interventions (NPIs) to curb transmission, including stay-at-home orders, the closure of schools and retail spaces, mandated face masks in public, and encouragement of social distancing and hand hygiene. Studies from Hong Kong, France, Japan, Singapore and Australia have shown these measures have also coincided with a decline in the number of cases of the seasonal respiratory viruses over the same time period, as compared to prior seasons [3-7]. A similar trend occurred in the U.S., with the number of people with influenza-like illness (ILI) [8] for the 2019-2020 season decreasing earlier than expected as compared to the 2015-2016, 2008-2009, and 2002-2003 influenza seasons [9].

In this study, we sought to define the impact of NPIs intended to curb SARS-CoV-2 transmission on the rates of infection from influenza A, influenza B and RSV in two large U.S academic centers located in Atlanta, Georgia and Boston, Massachusetts from January through May 2020. The heterologous impact of NPIs has important implications for clinicians and laboratories preparing for future respiratory virus seasons.

We conducted a retrospective review of medical records for all inpatient and outpatient adults who underwent testing for respiratory viruses over the previous 5 seasons (09/01/2015 – 05/30/2020), at the Emory Healthcare system (EHC) and associated clinics in Atlanta (n =46,575 people) and the Mass General Brigham (MGB) Healthcare System in Boston (n = 107,260 people). Each respiratory virus season was defined as beginning on September 01 (week 36) of a given year and extending through May 30 (week 22) of the following year. The weeks were assigned according to the International Organization for Standardization (ISO) week date standard, which defines the start and end of a week on Monday and Sunday, respectively.

Specimens were obtained using nasopharyngeal swabs and tested by the eSensor® Respiratory Viral Panel (GenMark Diagnostics, Inc., Carlsbad, CA) at EHC and the Cepheid Xpert or the Hologic Panther Fusion at MGB, per manufacturer instructions. A small subset of patients at MGB and EHC were tested for SARS-CoV-2 using oropharyngeal swabs and lower respiratory tract samples. Total cases of influenza A and B, RSV, and SARS-CoV-2 were calculated for each week of each season. These case counts were then used to calculate the pathogen-specific reproductive number over time, R_t_, given by the ratio of the number of new positive cases in one week to the number of positive cases in the previous week. A reproductive number greater than 1.0 corresponds to an increase in the number of cases between two consecutive weeks, whereas a value between zero and 1.0 corresponds to a decrease.

To evaluate the impact of NPIs, we compared the R_t_ for influenza A, influenza B, and RSV averaged over all previous seasons to the current season (2019-2020) using a bootstrapping approach. Specifically, we first drew randomly from a binomial distribution to generate absolute counts for each virus, for each week of each season. The binomial distributions were parameterized using the number of tests in a given week as the number of trials and the percent of tests positive for the virus as the success probability. Once values of absolute counts were drawn, the bootstrapped R_t_ value for that pair of consecutive weeks was generated by taking the ratio of the counts in one week to the counts in the previous week. This procedure was repeated to generate 1,000 independent samples. Mean R_t_ values were calculated for a given pair of consecutive weeks by averaging over a set of four R_t_ values drawn from the bootstrapped samples, representing one value from each of the previous four seasons (2015-2016 through 2018-2019). Similarly, 95% CIs were calculated from the distribution of R_t_ values generated by the bootstrapped samples for each week of each season. We utilized only the first positive test for each patient, to reduce bias introduced by patients with persistently positive tests. For the 2019-2020 season, mean R_t_ values and 95% CI were calculated. All analyses were conducted using R (v 4.0.0).

For influenza A infections in Atlanta, the effective reproductive number R_t_ dropped below 1.0 on week 12 of the 2019-2010 season, whereas in previous seasons the effective reproductive number did not consistently drop below 1.0 prior to week 15 (Figure 1). This decrease in influenza A cases coincides with the first detection of SARS-CoV-2 cases in Atlanta, which began in week 11, and coincides with the city of Atlanta restricting large gatherings, closing bars, and issuing a stay-at-home order (weeks 12-13). Case counts for influenza B fell to near zero cases per week after week 13, precluding the calculation of reproductive numbers. In prior years, R_t_ values decreased to <1.0 after week 15, which shows that rates of influenza B infections decreased earlier in the 2019-2020 season relative to past seasons. In Atlanta, RSV R_t_ values did not drop below 1.0 until week 13 during the current season and week 14 for previous seasons (Supplemental Figure 1).

**Figure 1:**
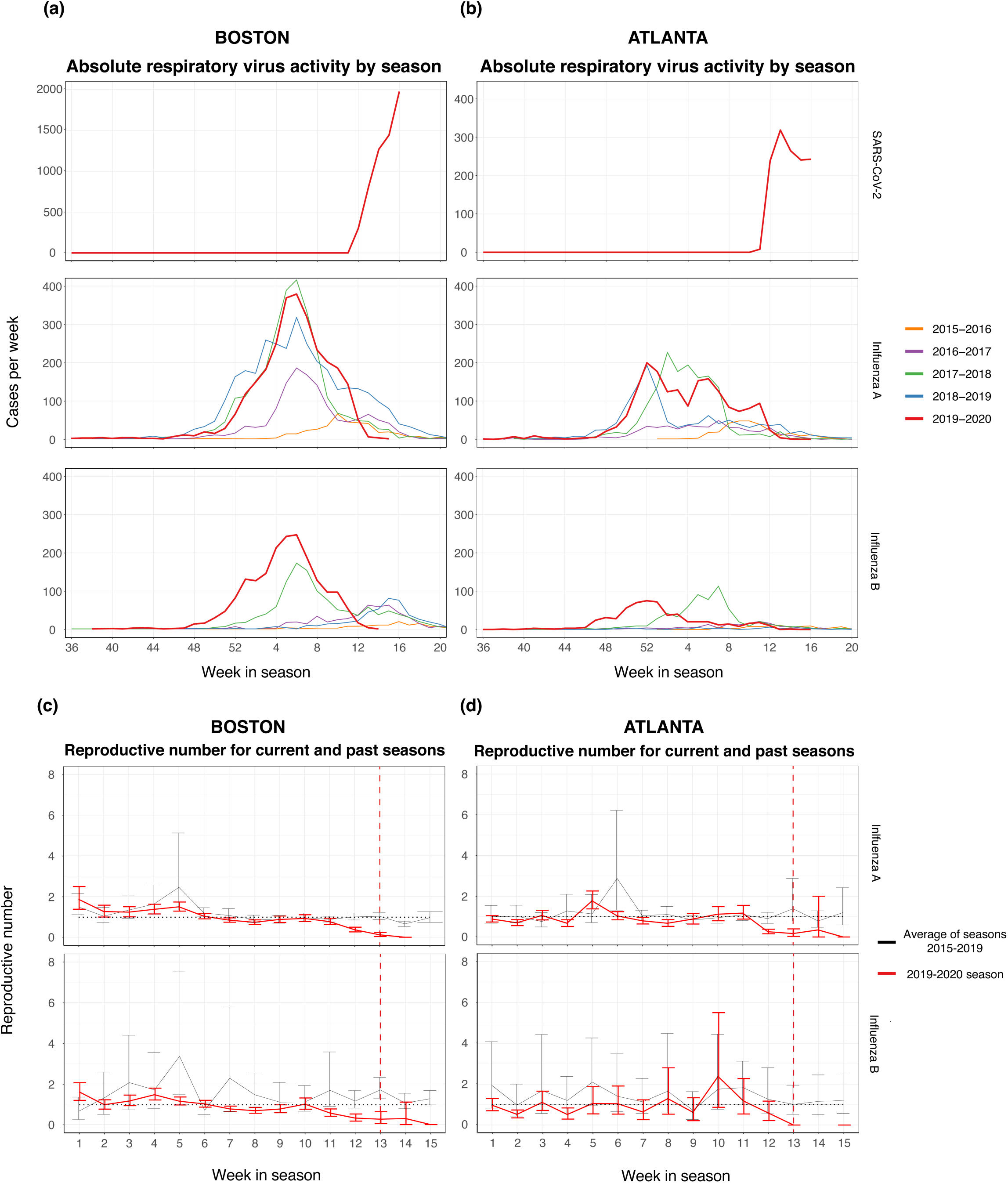
Cases per week of influenza A and B and SARS-CoV-2 of seasons 2015-2020 in the MGB Healthcare systems in Boston (**panel A**) and in the Emory University in Atlanta (**panel B**) by week of respiratory virus season (note the change in y-axis scale for Boston SARS-CoV-2 cases, panel A). Comparison of the reproductive number over time, R_t_, for influenza A and B in prior seasons (the average of the 2015-2016 through 2018-2019 seasons, as depicted by the black line) versus the 2019-2020 season as depicted by the red line, for Boston (**panel C**) and Atlanta (**panel D**). The black dotted horizontal line denotes R_t_ of 1.0. The red dotted vertical line represents the approximate date when NPIs were initiated in Atlanta and Boston (week 13).

Similar trends were observed in Boston (Figure 1). For influenza A infections in Boston, the effective reproductive number R_t_ dipped below 1.0 on week 7 and fell to 0 at week 14 of the 2019-2020 season. The effective R_t_ for influenza A in previous seasons did not remain below 1.0 before week 15. For influenza B, R_t_ was consistently below 1.0 at week 11 while the R_t_ of influenza B in past seasons did not consistently drop below 1.0 before week 15. In Boston, social gatherings were restricted to 25 people or less on week 11, and stay-at-home advisories were issued on week 13, which corresponds to the decrease of influenza A and B cases. For RSV, R_t_ predominantly fell below 1.0 from weeks 12-15 for the 2019-2020 season while previous seasons’ R_t_ values remained above 1.0.

The implementation of NPIs was a critical factor in reducing the spread of SARS-CoV-2 globally. We provide evidence from two geographically distinct metropolitan U.S. cities that NPIs may also have exerted a collateral beneficial effect on influenza A and B and RSV by virtue of reducing opportunities for transmission. Although the overall case counts of SARS-CoV-2 and the response to the pandemic differed in Atlanta and Boston, the results demonstrate the importance of social distancing measures. These findings should help inform pre- and post-test probabilities when evaluating patients presenting with symptoms consistent with an upper or lower respiratory tract viral syndrome, as well as the interpretation of respiratory virus panel testing performed for asymptomatic screening purposes.

The dynamics of influenza, RSV, and SARS-CoV-2 epidemiology are complex, and the interplay between these respiratory viruses will become increasingly important to dissect as a new influenza season emerges in the fall of 2020. Globally, the effects of decreased influenza cases from January to May 2020, due to either decreased transmission or decreased surveillance, may confound the ability to accurately predict which strains will be circulating in the fall. With decreased international travel and border restrictions, there may also be a change in the temporal dynamics of influenza illness in 2020-2021, and the circulating strains may differ from those predicted in February 2020. Further study is warranted to determine the downstream effects that SARS-CoV-2 will have on influenza transmission and epidemiological trends.

Limitations which must be acknowledged include the possibility of decreased reporting of influenza and RSV in March due to a decline in overall outpatient clinic visits (Supplemental Figure 2, total influenza and RSV tests conducted), preservation of viral media and equipment for SARS-CoV-2 testing, and transitions in hospitals to prioritize SARS-CoV-2 testing over other viral diagnostics for hospitalized patients [10]. Complex viral factors and dynamics between circulating SARS-CoV-2 and influenza viruses may have also influenced this trend. The phenomenon of respiratory virus “interference” has been described for epidemics of influenza and other respiratory viruses, in which one epidemic delays the start or accelerates the end of the other viral epidemic [11-13]. The underlying immune mechanisms for virus interference are not well elucidated but may be due to temporary non-specific immunity for a given acute respiratory infection [14].

In summary, the data demonstrate that NPIs are likely effective in reducing both influenza and SARS-CoV-2 transmission. Concurrent epidemics of influenza and SARS-CoV-2 have the potential to result in diagnostic confusion, and significant morbidity and mortality and could overwhelm the healthcare system. Consequently, NPIs, in addition to widespread utilization and uptake of influenza vaccination, will be important public health tools to deploy to mitigate the threat of these respiratory viruses. Ongoing diagnostic and surveillance efforts for influenza, RSV and SARS-CoV-2 must be maintained to further explore the transmission mechanisms and interplay between these respiratory viruses.

## Data Availability

Datasets (de-identified) can be made available on reasonable request to the corresponding author

## Conflict of Interest

The authors report no conflict of interest.

## Funding

No funding to report.

## Acknowledgments

We would like to acknowledge our laboratory colleagues at the Emory University Healthcare and Mass General Brigham Healthcare System Microbiology and Molecular laboratories and who have worked tirelessly to provide necessary care to our patients during this time.

## Figure Legends

**Supplemental Figure 1:**
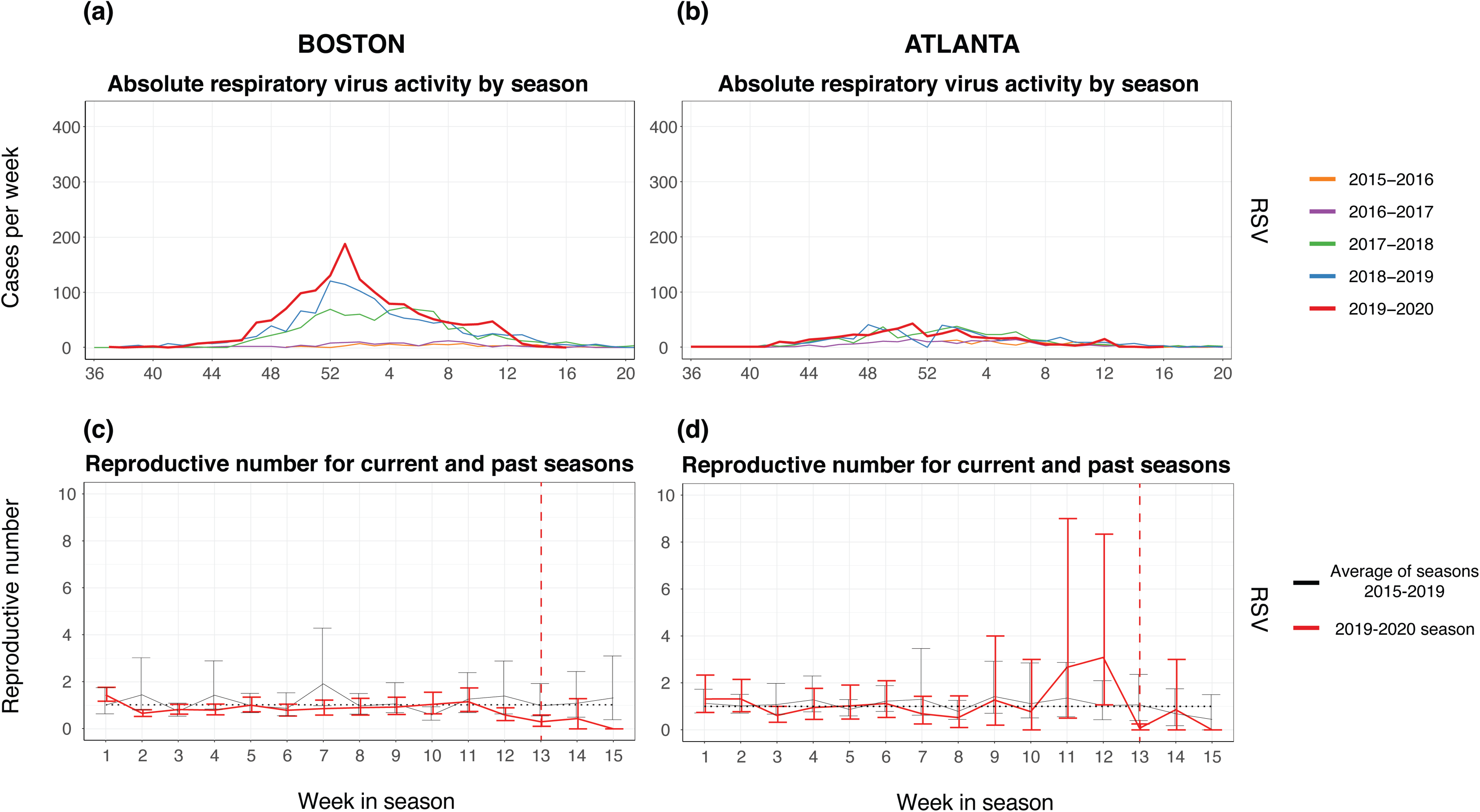
Cases per week of RSV of seasons 2015-2020 in the MGB Healthcare systems in Boston **(panel A)** and in the Emory University in Atlanta **(panel B)** by week of respiratory virus season. Comparison of the reproductive number over time, R_t_, for RSV in prior seasons (the average of the 2015-2016 through 2018-2019 seasons, as depicted by the black line) versus the 2019-2020 season as depicted by the red line, for Boston **(panel C)** and Atlanta **(panel D)**. The black dotted horizontal line denotes R_t_ of 1.0. The red dotted vertical line represent the approximate date when NPis were initiated in Atlanta and Boston (week 13).

**Supplemental Figure 2:**
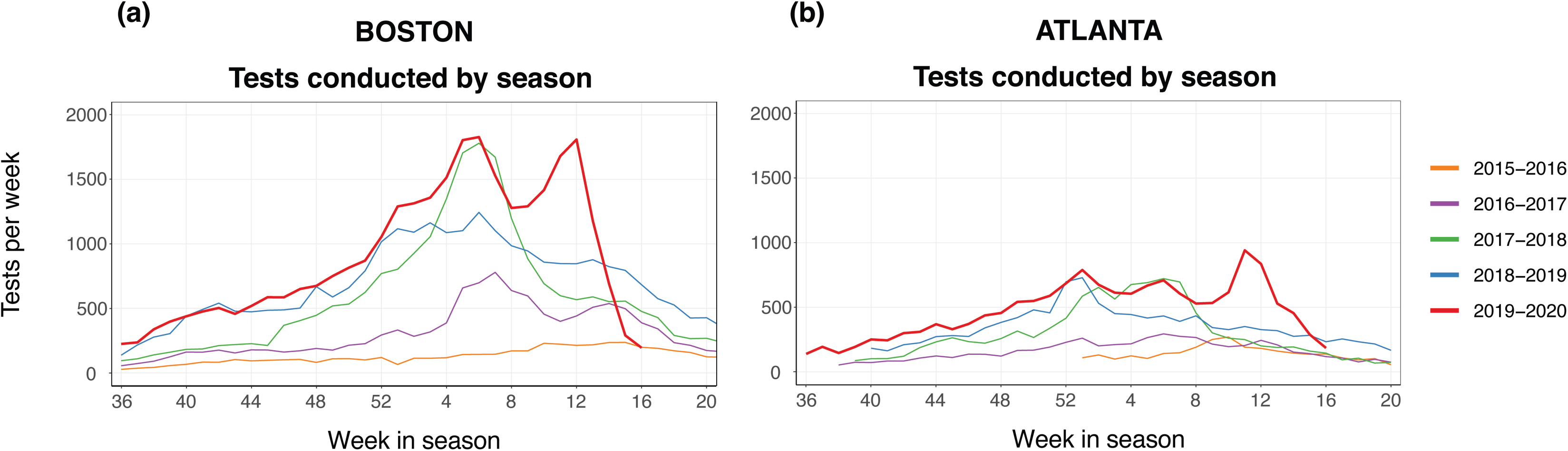
Total tests conducted for influenza A and B combined for seasons 2015-2020 in the MGB Healthcare systems in Boston **(panel A)** and in the Emory University in Atlanta **(panel B)** by week of respiratory virus season.

## Notes

### Competing Interest Statement

The authors have declared no competing interest.

### Author Declarations

The study was approved by IRB bodies at each respective institution (Emory University and Mass General Brigham)

